# Origin, pattern of bone metastasis and need for surgery in patients with Metastatic bone disease treated at tertiary hospital in Northern Tanzania. A hospital based cross sectional study

**DOI:** 10.1101/2024.02.21.24303155

**Authors:** Mathias Ncheye, Elifuraha Maya, Furaha Serventi, Faiton Mandari, Rogers Temu, Peter Magembe, Honest Massawe

## Abstract

Bones are the third most common sites for cancer metastasis after lung and liver. Bone metastasis cause skeletal complications (SRE’s) that affect the quality of life of patients with bone metastasis. Management of these patients depend on the primary tumor and pattern of bone metastasis.

The aim of this study was to describe the origin, distribution pattern of bone metastases, common SRE’s and proportion of patients with bone metastasis that need surgery.

A cross sectional study was conducted among cancer patients with Metastatic bone disease attending KCMC Hospital from November 2022 to April 2023. Data was collected from patient’s files, histopathology and radiology reports by a structured extraction sheet. VAS, SINS and Mirel’s scores was used to document pain severity, spinal instability and fracture risk respectively. The ASIA impairment scale was used for documenting neurological deficits in patients with spine metastasis. Data was entered and analyzed in SPSS version 25.0

A total of 72 participants were enrolled. Their mean age was 69 ± 11 years and (75%) were male. Prostate cancer (65.3%) was the leading cause of metastatic bone disease followed by breast cancer (18.1%). Most of patients with MBD have multiple lesions (91.7%) involving multiple sites but the spine (93.1%) was the mostly affected site. Osteoblastic lesions were the predominant radiological type by 59.7% followed by osteolytic lesions which accounted for 23.6% of the study participants. 30.6% had pathological fractures and half of these occurred in patients with osteolytics lesions. 36.1% of the study participants had an indication for surgical treatment of the bone metastasis.

Most of MBD originate from Prostate and Breast cancer giving multiple lesions involving multiple sites but the spine remains to be the most affected site. Even though about a third of the patients had SRE’s that needed surgical intervention but few are expected to be operated considering the prognosis. This calls for more emphasis on prevention of SRE’s and use of appropriate less invasive therapies to prevent progression of the disease.

## Introduction

In the US, it is estimated that about 5% of all cancer patients have bone metastasis, the common primary cancers been breast, prostate and lung cancer accounting for almost 70% of all bone metastasis. (1,2)

In our setting, Bone metastasis is one of the most common musculoskeletal tumors just second to osteosarcoma.(3) Studies that have investigated on common primary malignancies like breast and prostate cancer have shown that most patients present late and bone metastasis is present in 24-58% of the patients.(4–7)

Most patients present to the orthopedic surgeon with pain, pathological fractures or symptoms due to spinal cord compression. These are called skeletal related events (SRE’s). These cause significant morbidity and affect the quality of life of patients with cancer. The onset of SRE’s has been shown to vary depending on the primary tumor, metastatic site and the radiological pattern.(8,9)

Surgical management of bone metastasis has been shown to help improve quality of life, in patients with metastatic bone disease. It can be done to fix a pathological fracture, to decompress spinal cord compression due to spine metastasis or even to prevent an impending pathological fracture in long bones. Surgery is also done for patients with intractable pain that does not respond to opioids or radiotherapy. (10–12)

Due to the advancement of cancer treatment now patients with cancer live longer and so the incidence and prevalence of bone metastasis is also Increasing. The data on on the distribution pattern of bone metastasis and the proportion of patients with metastatic bone disease who need surgery in our setting is limited.

We aim to describe the origin, distribution pattern of bone metastases and proportion of patients with bone metastasis that need surgery.

## Materials and Methods

### Study Area and setting

We conducted a Hospital based cross section study among cancer patients with metastatic bone disease attending at Kilimanjaro Christian Medical center (KCMC), a tertiary and consultant hospital in Northern Tanzania with bed capacity of 721. It serves approximately 11 million people from this part of the country as per 2022 census.

Study participants were recruited at the clinic of cancer care center after identifying them in the cancer registry. Only those with confirmed bone metastasis and who were receiving care at the hospital from 1^st^ November, 2022 to 30^th^ April, 2023 were included. Data was collected during this 6 Months period but data processing and analysis was done from May 2023 to July 2023. The author had access to identification informations of the participants during and after data collection.

### Eligibility criteria

#### Inclusion criteria

All cancer patients with metastatic bone disease treated at KCMC from November 2022 to April 2023.

#### Exclusion criteria

Patients who have already received surgical treatment of bone metastasis, no histological results of primary tumor and those with missing information were excluded.

### Variables

Outcomes were skeletal related events (SRE’s) and Need for surgery. Predictors were Age, Sex, Type of primary cancer, Location of metastatic lesion, number of lesions and radiological type of metastatic lesion.

### Data sources and measurements

Demographic data was obtained by a structured questionnaire after obtaining a signed informed consent (S1 File) and secondary data was collected from patient’s files, histopathology and radiology reports by a structured extraction sheet. Skeletal-survery CT scan and MRI images were reviewed by the principle investigator to document the pattern and presence of spinal cord compression respectively.

VAS, SINS and Mirel’s scores was used to document pain severity, spinal instability and long bone fracture risk respectively. These scores have been shown to be valid and reliable than clinical judgment.(13–15) The ASIA impairment scale (S2 File) was used for documenting neurological deficits in patients with spine metastasis.

### Study size

433 cancer patients were registered during the study period. A total of 130 patients had metastatic cancer at different sites, these were filtered and we were able to identify a total of 79 patients with metastatic bone disease. 7 were excluded due to missing information. 72 study participants were left for the final analysis.

### Data Processing and Analysis plan

Data was entered and analysed by SPSS version 23.0 statistical package. Categorical variables were summarized by frequencies and percentages in tables, bars and charts. Numerical variables were summarized by measures of central tendacyi.e mean (S.D) and median (Range)

Chi square/fishers exact test was performed to compare differences in proportions of SRE’s between different groups and P value less than 0.05 was considered statistically significant.

### Ethics statement

Approval to conduct the research was obtained from the Kilimanjaro Christian Medical University College Research, Ethics and Review Committee (CRERC) with clearance number PG94/2022. Written informed consent was obtained from participants before participating in the study.

## Results

### Social demographic information and other patients’ Characteristics N=72

Most of patients with metastatic bone disease were elderly with a mean age was 69±11 years and the majority (47.1%) were aged between 60 to 74 years. 75% of the study participants were male and 76.4% of the study participants had some form of health insurance.

More than half of the study participants were on chemotherapy (59.7%) and/or hormonal therapy (58.3%). 43.1% of the participants had received surgical treatment for resection of the primary tumor and 38.9% were on bisphosphonates. Radiotherapy was the least among the treatments with only 11.1% of the participants who had received it during the course of their treatment. This information is shown in tables 1 and 2 below

**Table 1.** Social demographic characteristics N=72.

| Variable | N (%) |
| --- | --- |
| **Age (Years)** |  |
| $\leq 59$ | 13(18.1) |
| 60-74 | 34(47.1) |
| 75+ | 25(34.7) |
| **Mean (S.D) = 69 (11) Years** |  |
| **Gender** |  |
| Male | 54(75.0) |
| Female | 18(25.0) |
| **Insurance status** |  |
| Insured | 55(76.4) |
| Not Insured | 17(23.6) |
| **Total** | **72(100.0)** |

**Table 2.** Type of treatment that the study participants were receiving.

| **Treatment** | **Received**<br>**N (%)** | **Not received**<br>**N (%)** |
| --- | --- | --- |
| Surgical | 31(43.1) | 41(56.9) |
| Hormonal | 42(58.3) | 30(41.7) |
| Chemotherapy | 43(59.7) | 29(40.3) |
| Radiotherapy | 8(11.1) | 64(88.9) |
| Bisphosphonates | 28(38.9) | 44(61.1) |

### The commonest primary cancers causing metastatic bone disease

Prostate cancer was found to be the leading cause of metastatic bone disease accounting for 65.3% followed by breast cancer 18.1%, Lung cancer 4.2%, Colorectal Cancer 4.2%. This information is summarized in table 3 below.

**Table 3.** The Common primary cancers causing metastatic bone disease.

| **Primary** | **N (%)** |
| --- | --- |
| Breast Cancer | 13(18.1) |
| Bronchus and lung cancer | 3(4.2) |
| Colorectal cancer | 3(4.2) |
| Gastric adenocarcinoma | 1(1.4) |
| Liver cancer | 1(1.4) |
| Ovary cancer | 1(1.4) |
| Prostate adenocarcinoma | 47(65.3) |
| Thyroid adenocarcinoma | 2(2.8) |
| Unknown | 1(1.4) |
| **Total** | **72(100.0)** |

### Pattern of bone metastasis among patients with metastatic bone disease

Most of the study participants (91.7%) had multiple metastatic bone lesions involving more than one site. The spine was involved in 67(93.1%) participants followed by the pelvis 38 (52.8%) and long bones 19 (26.4%). Other bones involved were the rib cage, scapula, clavicles, sternum, mandibles and skull which together accounted for 38.9%. This information is shown in Table 4 below

**Table 4.** Location of bone metastasis.

| Location of bone metastasis | N (%) |
| --- | --- |
| Spine | 67(93.1) |
| Pelvis | 38(52.8) |
| Femur | 15(20.8) |
| Humerus | 3(4.2) |
| Other*** | 29(40.3) |

\*\*\*Other include ribcage, Thoracic, Scapula, Skull, Tibia, Clavicle and Mandible sterunum

#### 4.2.1 **Radiological pattern**

We found that majority 59.7% of the study participants had osteoblastic/sclerotic while 23.6% had osteolytic and 12 (16.7%) had mixed sclerotic and lytic lesions. This information is summarized in table 5

**Table 5.**
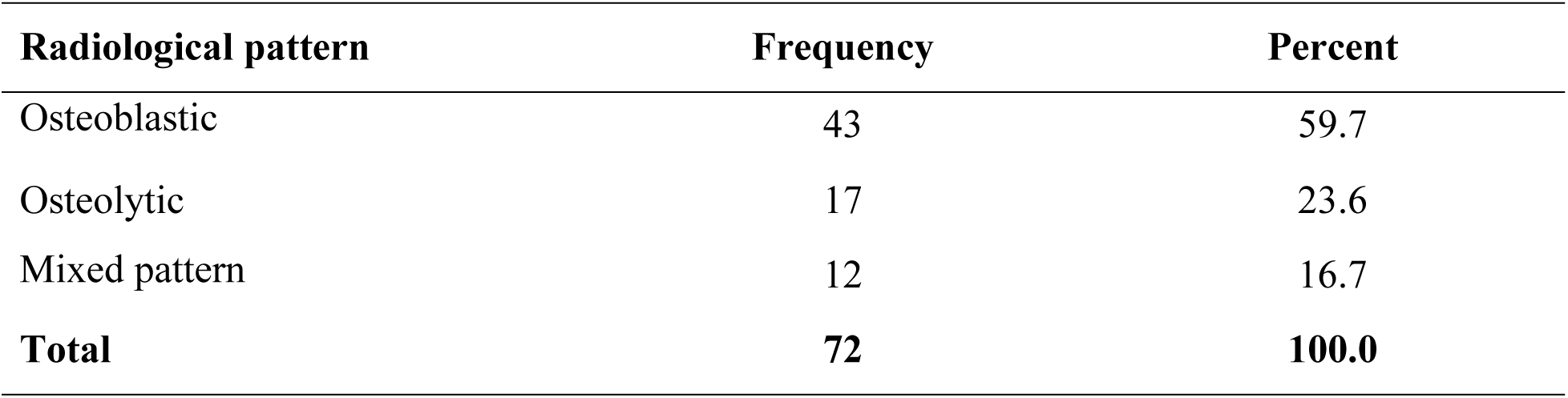
Distribution of study participants based on radiological pattern of the metastatis bone lesions.

### 4.3 Skeletal related Events and Need for surgery

41.7% of the study participants had functional pain requiring opioid analgesics (Fig. 1) and 31% had pathological fractures. 50% of the pathological fractures occurred in patients with osteolytic lesions (p – value < 0.001) while only 27.3% occurred in patients with osteoblastic lesions. The rest of the fractures occurred in patients with mixed type of lesions. Table 6 shows the distribution of the pathological fractures by Age, Gender, type of primary cancer, radiological pattern, number of lesions and site of metastasis.

**Fig 1.**
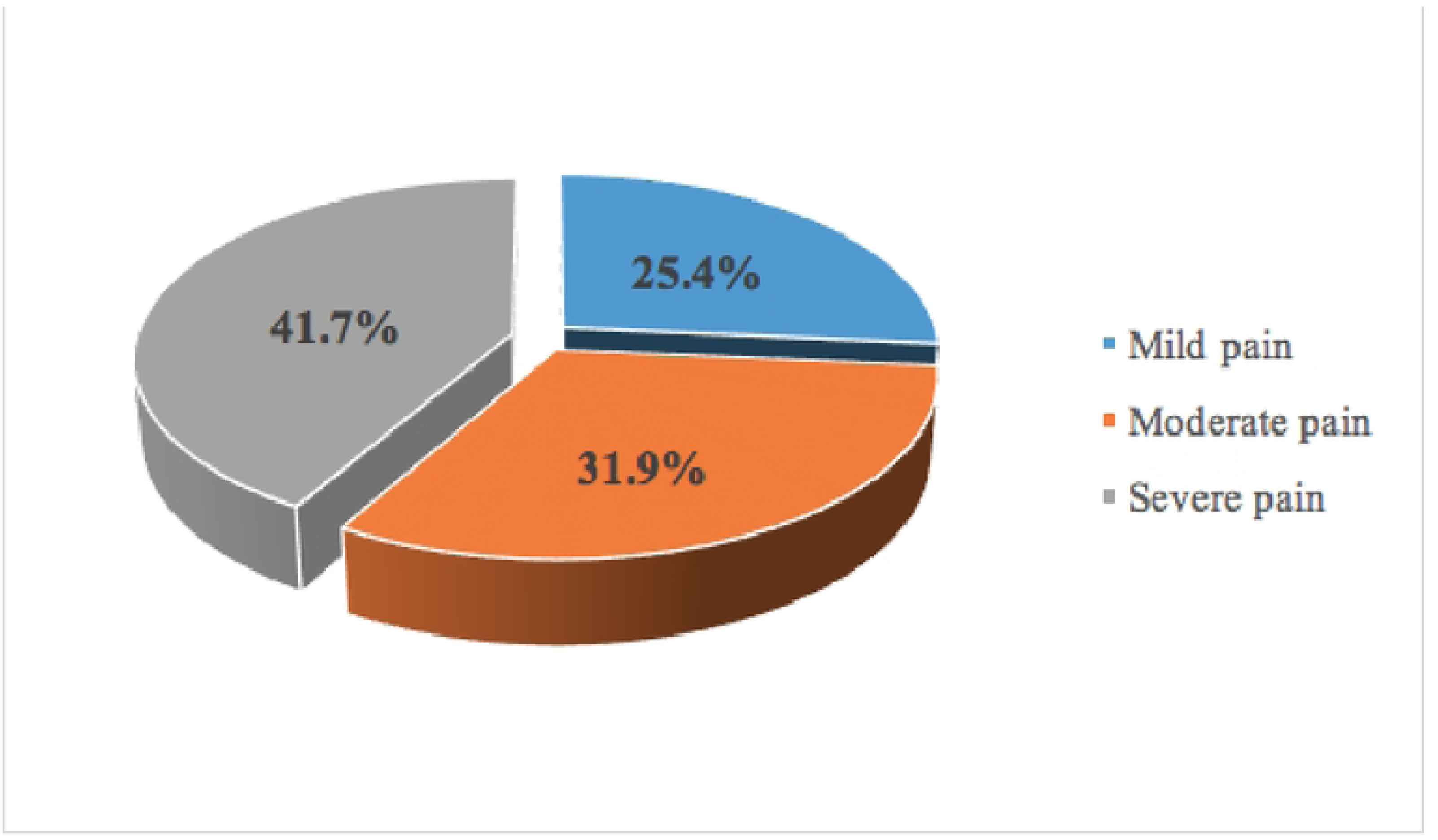
proportions of participants based on their pain severity.

**Table 6.** Pathological fractures according to age, sex, type of primary cancer and pattern of bone metastases (fisher’s exact test)

| **Independent variable** | **pathological fractures** |  | **Total** | **P-value** |
| --- | --- | --- | --- | --- |
|  | **Yes** | **No** |  |  |
| **Age** |  |  |  |  |
| <=59 | 8(36.4) | 5(10.0) | 13(18.1) | 0.032 |
| 60-74 | 9(40.9) | 25(50.0) | 34(47.2) |  |
| 75+ | 5(22.7) | 20(40.0) | 25(34.7) |  |
| **Gender** |  |  |  |  |
| Male | 12(54.5) | 42(84.0) | 54(75.0) | 0.016 |
| Female | 10(45.5) | 8(16.0) | 18(25.0) |  |
| **Primary cancer** |  |  |  |  |
| Prostate Cancer | 9(40.9) | 38(76.0) | 47(65.3) | 0.015 |
| Breast Cancer | 7(31.8) | 6(12.0) | 13(18.1) |  |
| Others | 6(27.3) | 6(12.0) | 12(16.7) |  |
| **Radiological pattern** |  |  |  |  |
| Osteoblastic | 6(27.3) | 37(74.0) | 43(59.7) | <0.0001 |
| Osteolytic | 11(50.0) | 6(12.0) | 17(23.6) |  |
| Mixed pattern | 5(22.7) | 7(14.0) | 12(16.7) |  |
| **Number of lesion** |  |  |  |  |
| Single/Solitary | 0(0.0) | 6(12.0) | 6(8.3) | 0.168 |
| Multiple | 22(100.0) | 44(88.0) | 66(91.7) |  |
| **Site** |  |  |  |  |
| Spine | 16(72.7) | 51(79.7) | 67(77.9) | 0.163 |
| Long bones | 06(27.3) | 13(10.3) | 19(22.1) |  |

Among 67 patients with metastases to the spine, 23.9% had pathological fractures, 6% patients had spinal cord compression, 13.4% had spine instability while 70.1% had potential instability (Table 7).

**Table 7.**
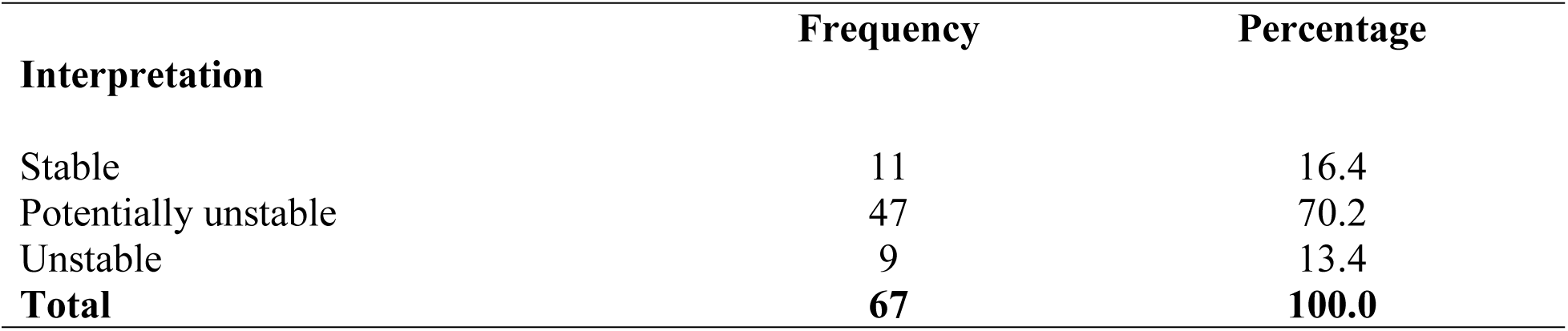
Spine instability based on SINS score.

| Interpretation | Frequency | Percentage |
| --- | --- | --- |
| Stable | 11 | 16.4 |
| Potentially unstable | 47 | 70.2 |
| Unstable | 9 | 13.4 |
| **Total** | **67** | **100.0** |

We found 17 patients with long bone metastases and a total of 19 long bones were involved. Among the 19 long bones, 06 (31.6%) had pathological fractures and 07 (36.8%) had impending fracture requiring prophylactic fixation. A total of 26 (36.1%) study participants had absolute indications for surgical intervention due to metastatic bone disease. These Indications with the corresponding number of study participants is shown in table 8.

**Table 8.**
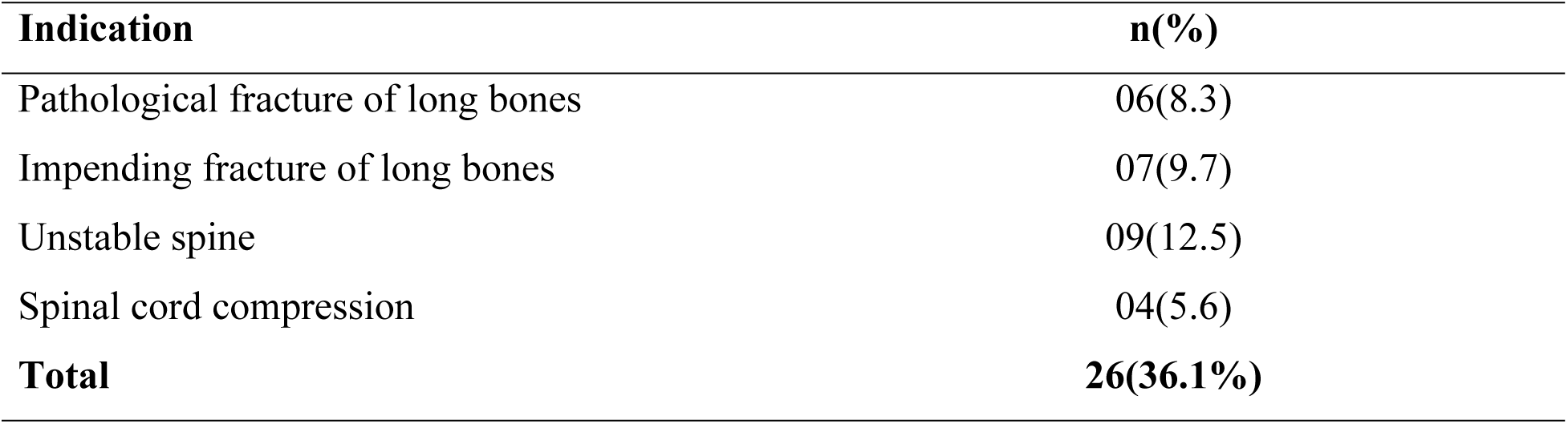
Proportion of study participants with absolute indications for surgical intervention of MBD.

| Indication | n(%) |
| --- | --- |
| Pathological fracture of long bones | 06(8.3) |
| Impending fracture of long bones | 07(9.7) |
| Unstable spine | 09(12.5) |
| Spinal cord compression | 04(5.6) |
| **Total** | **26(36.1%)** |

## Discussion

### Discussion of key findings

#### Characteristics of the study participants

In our study we found that most of patients with metastatic bone disease were elderly with a mean age of 69 +/- 11 years. These findings are similar to a study done in Nigeria among patients with metastatic prostate cancer where mean age was 67+/-1.8 years. (16) They are consistent with other studies which also showed that increasing age is associated with more skeletal metastasis. (5,7)

Men accounted for 75% of the study participants which is reflected by the large proportion of patients in this study having prostate cancer as the primary cancer leading to bone metastases.

76.4% of the study participants had some form of health insurance which is a good observation considering the cost of treatment in patients with metastatic bone disease.

More than half of the study participants were on either chemotherapy and/or hormonal therapy with bisphosphonates. Few had received radiotherapy at a peripheral center because it is currently not offered at KCMC Hospital and most of the patients who need RT are usually reffered to a center where it is offered. Efforts are being made to start providing Radiotherapy services at KCMC and RT bankers are under construction as we write this report.

The management of patients with bone metastases needs a multidisciplinary team approach. The main goal is for symptom palliation and prevention of skeletal related events thus improve survival and the quality of life among these patients. Conventional cytotoxic chemotherapy has little effect on bone as compared to bone targeting therapies which have direct effect on bone remodeling. The use of bone targeting therapies has paved way to achieving this goal and are divided into local regional and systemic therapies. (8,10)

Local regional therapies include orthopedic surgery and radiotherapy which are aimed at pain relief and management of skeletal related events such as pathological fractures and spinal cord compression. Systemic bone targeting therapies include the use of Bone resorption inhibitors (BRI’s). Bisphosphonates is one example of the commonly used inhibitors of bone resorption.

Bisphosphonates have also been shown to stimulate innate anti-cancer immune response by up regulating γδT-cells. (8)

Denosumab is another bone resorption inhibitor which is a monoclonal antibody against RANK-L. It reduses osteoclast activity by impairing the activation of osteoclasts. It also causes bone remodeling and increase survival of patients with bone metastases. Donosumab has been shown to be superior to zolendronic acid in reducing the likelihood of pathological fractures and other SRE’s but there was no significant difference in overall survival improvement between denosumab and zolendronic acid. (9,17,18)

Denosumab was also shown to have less adverse events than bisphosphonates. The side effects of bisphosphonates include osteonecrosis of the Jaw, gastro-intestinal upset and gastritis, hypo-calcemia, fevers and skin rash.

Other systemic bone targeting therapies under study include the use of Tyrosine Kinase Inhibitors and Immune Checkpoint Inhibitors but these are out of scope of this study.(8)

#### Common primary Cancers causing Metastatic Bone Disease

Prostate cancer was found to be the leading cause of metastatic bone disease followed by breast cancer, Lung cancer, Colorectal Cancer. In our setting more than half of patients with prostate and Breast cancer have been shown to present with bone metastasis metastasis. (7)(5) These cancers have also been shown to be the commonest primary cancers in studies done in other parts of the world. (1,2,9,19,20)

This is because the establishment of bone metastasis involve an intricate relationship between the primary tumor and the bone micro-environment. It involves phases from the detachment of tumor cells from the primaries to the colonization of the metastatic site.

The vertebral venous plexus is a system of valve less veins that drains the chest cavity and pelvis. It was postulated by Batson in 1940 that this system of veins is responsible for the spreading of cancer cells to the bones. However, this does not explain the preferential homing of cancer cells in bones. (8,21)

Paget hypothesized what is called the “Seed and soil model” where the tumor cells are the seeds that will flourish and grow in a micro-environment of the organ that will provide a suitable soil. The bone provides a favorable microenvironment for tumor growth due to the presence of calcium, hypoxia, acidosis and various growth factors that are released from the mineralized bone matrix. (8,21)

The homage of tumor cells into the bones is influenced by various integrins and chemokines such as CCXR4 that help the tumor cells attach to the bone marrow endothelium.

Metastatic bone tumors thrive in the bone micro-environment by a feed forward vicious cycle. This cycle involves an interplay between the tumor cells, osteoclasts, osteoblasts, cytokines and growth factors.

#### Pattern of Bone metastases

In most of the study participants, more than one site was involved and almost all had multiple metastatic lesions (Fig. 2). This pattern was seen to be common in studies done in other parts of the world (16,20,22)

**Fig 2.**
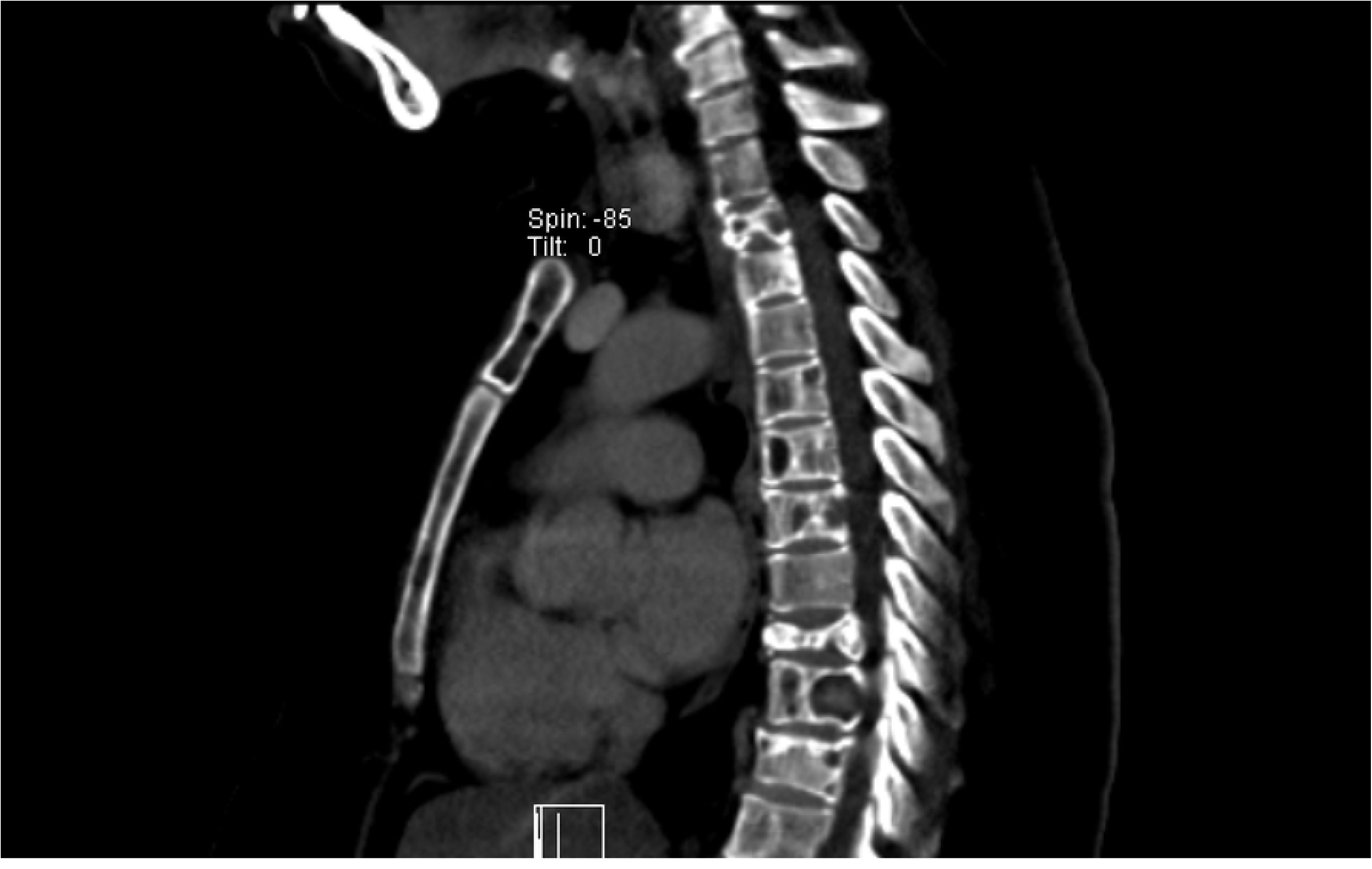
Multiple lesions of the spine; Sagittal CT images of the thoraco-lumbar spine of a lady who had breast cancer showing multiple metastatic lesions on the spine with pathological fracture of vertebral body

The spine was involved in 93.1% participants followed by the pelvis 52.8% and long bones 26.4%. Other bones involved were the rib cage, scapula, clavicles, sternum, mandibles and skull which together accounted for 38.9%. These findings are similar to other studies where the spine was shown to be the preferred metastatic site. (16,22,23)

These findings are explained by the hematogenous spread of the cancer cells through the Batson Venous plexus, thus increasing their propensity to lodge in the vertebral bodies.

Radiologically, 59.7% of the metastatic bone lesions were osteoblastic/sclerotic (fig. 3) while 23.6% were osteolytic and 16.7% had mixed sclerotic and lytic lesions. These findings are contrary to most of the studies we reviewed where osteolytic lesions were more common than osteoblastic lesions.(20,24,25) This might be explained by the fact that in their studies the commonest primary cancers are breast and lung malignancy while in our study the commonest primary is prostate carcinoma. Breast and lung cancer have been shown to form more lytic bone lesions than osteoblastic while prostate carcinoma forms more osteoblastic bone lesions.

**Fig 3.**
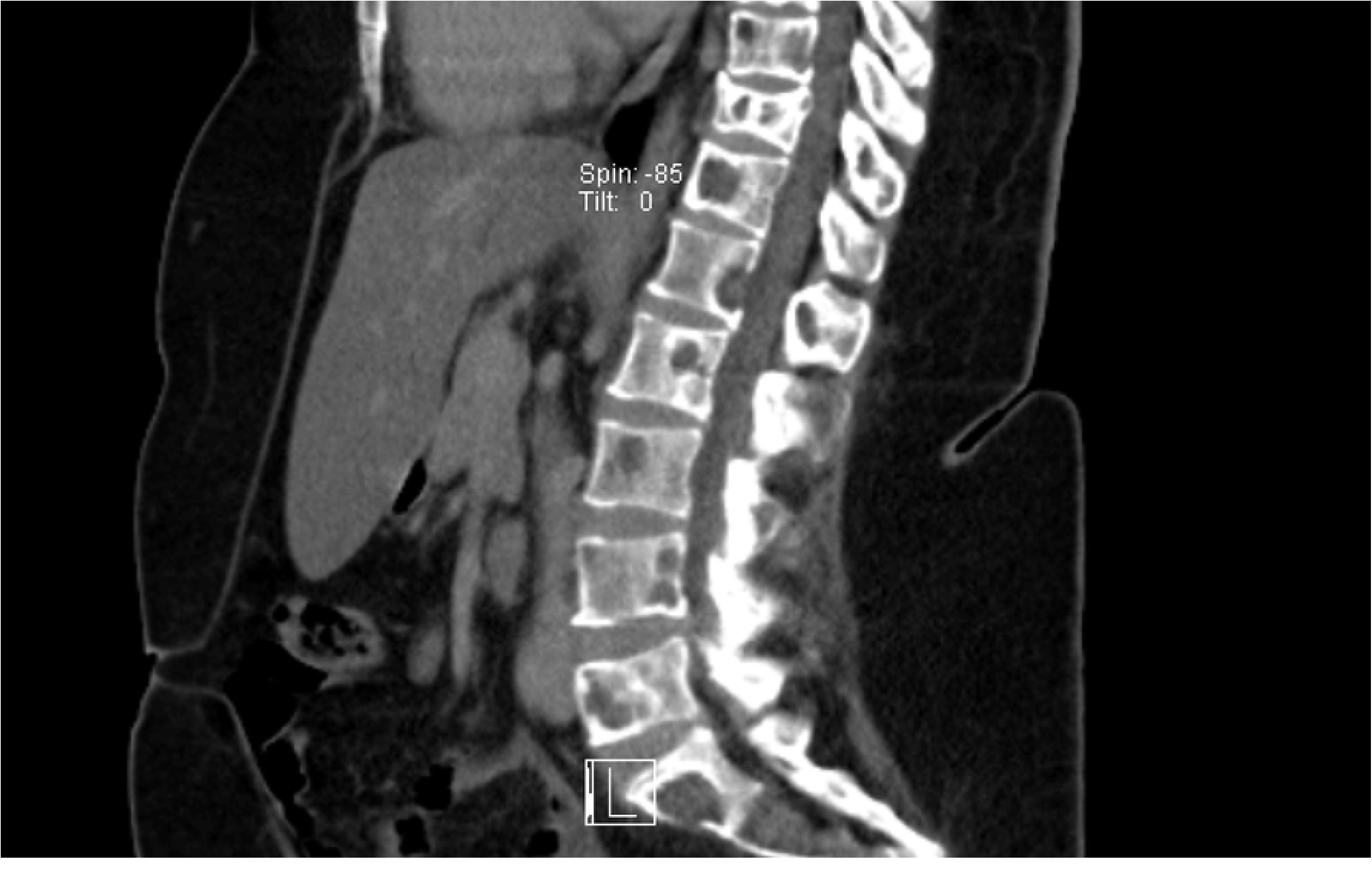
Osteoblastic lesions; X ray image showing diffuse osteoblastic lesions involving the pelvis and proximal femur in a patient with prostate carcinoma attending at our center

#### Skeletal related events and Need for surgery

About a third of the study participants had pathological fractures either on spine vertebrae or the long bones. Among the patients with metastases to the spine majority (70.1%) had potential instability, 23.9% had pathological fractures, 6% had spinal cord compression and 13.4% had spine instability. This rates are lower compared to studies done in other parts of the world where more than half of their study participants had pathological fractures and neurological deficits.(9,11) This might be explained by the larger proportion of our study participants having prostate carcinoma while in their studies the leading primary cancers were Breast and lung cancer.

Irrespective of the site involved, 41.7% of the study participants had severe pain requiring opioid analgesics. This finding differed with studies we reviewed where the value was higher in one study and very low in another. (11,24)

The onset of SRE’s has been shown to vary depending on the primary tumor, metastatic site and the radiological pattern. Tumors that form osteolytic metastatic lesions tend to cause more SREs such as pain and pathological fractures than those that form sclerotic lesions.(D’Oronzo*et al.*, 2019; Turpin and Duterque-coquillaud, 2020). In our study, half of the pathological fractures occurred in patients with osteolytic lesions and another 22.7% in mixed lytic and blastic lesions. This emphasizes on the risk of pathological fractures in patients with osteolytic lesions (fig 4).

**Fig 4.**
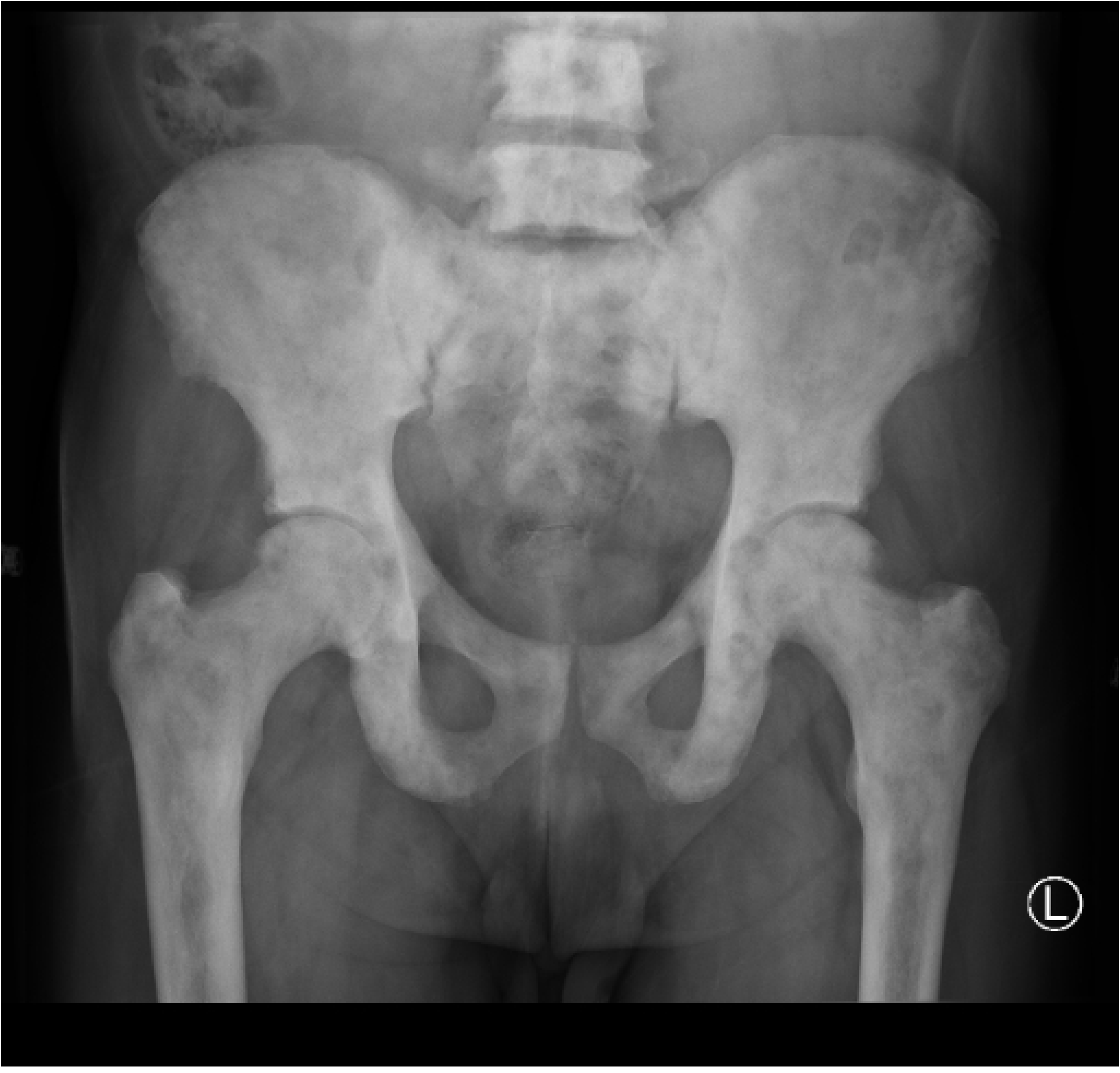
Osteolytic lesions; X ray images in a lady who had impending fracture of the femur (A) which was fixed by intramedullary nailing and then later presented with metastatic osteolytic lesion involving the shaft of the Tibia (B & C).

Hoban et al did a Retrospective cohort study to assess the fracture risk in patients with bone metastasis of the upper limb and found the overall fracture rate to be 76% after a mean follow up of 3.6 years. They also found out that a Mirels’ score of 7 or above had a high predictive value for fracture risk with higher sensitivity and specificity than that recommended in lower limb lesions. (26)

36.1% of the study participants had absolute indications for surgery. More than half of the patients with long bone metastases had either a pathological fracture or had an impending fracture requiring prophylactic fixation. This does not mean all these patients are fit for surgery and will end up being operated. Other important surgical considerations include assessment of the general medical condition of the patient such as the performance status, presence of other metastatic sites, expected survival and magnitude of the surgery to be performed. All these affect the decision on whether to operate or not to operate. The choice of surgery whether to do internal fixation or endo-prosthetic reconstruction should be chosen on the basis of the location of the lesion, the extent of bone destruction and the stability of the construct to outlast the life expectancy of the patient. (11,19,27)

In a Multi-Center Prospective study done in France that involved 245 patients to compare those treated by surgery for fracture fixation versus those who had surgery for prophylactic fixation, they found out that more than half of the patients were operated for fracture fixation. In this study they also found out that advanced age, VAS pain score > 6, WHO grade performance and upper limb location were independent predictors for surgical fixation.(25)

In the systematic review done by Errani to assess the treatment of long bone metastasis, patients with metastasis to the LL were operated more than those with metastasis to the upper limbs. In their study, patients were operated if expected survival was more than 6 weeks. (19)

Considering these factors, the need for surgery in patients with Metastatic bone disease further narrows down to few patients with longer anticipated survival post-surgery and those who are in good general medical condition.

This means that a huge proportion of the study participants will end up on other forms of treatment for MBD like Radiotherapy and bisphosphonates for pain relief, prevention of SRE’s and progression of the disease.

### Study Limitations and Strengths

#### Study limitations

Currently we do not have PET – CT scan and Radiotherapy services at our center. It is very likely that some lesions were missed and this was an important imaging modality for appropriate description of the pattern of the skeletal metastases. Also the lack RT services leads poor continuity of care as there is a significant number of patients who are reffered to other centers for Radiotherapy. This posed a challenge in acquiring information because it is not easy to access investigation results and data from a different center.

This is a single center observational study, so results can only be generalized with caution.

#### Strength

This study is first of its kind in our setting, we hope it will pave way for further studies on MBD

### Conclusion and recommendations

As stated prostate cancer is the leading cause of metastatic bone disease followed by breast cancer. So the role of targeted hormonal therapies cannot be over-emphasized. Also routine screening for breast cancer in females will help early detection and treatment to prevent advanced disease.

Most MBD present with multiple lesions involving multiple sites but the spine was the mostly affected site with a third of the patients presenting with pathological fractures. Despite this, a lesser number of the patients will be fit for surgery and so Radiotherapy and other non-surgical treatments will be preferred. A multicenter prospective study which will also look on survival and life expectancy will give more powerful findings that may guide in creating local protocols and guidelines for optimal treatment of these patients in our setting.

## Data Availability

All the data is available at the Cancer Care center registry of the Kilimanjaro Christian Medical center Hospital. This data cannot be accessed online for patient's privacy and ethical reasons

## Acknowledgements

I would like to thank the management of Kilimanjaro Christian Medical Centre for allowing me to conduct this study. I also extend my sincere gratitude to everyone in the Department of Orthopedics and Trauma at KCMC for their valuable contributions.

Special thanks to the staff at the Cancer Care center at KCMC with special note to Yotham Gwanika for his assistance during analysis and data processing from the cancer registry.

## Supporting information

S1 File. Informed consent

S2 file. ASIA reference manual

S3 file. STROBE statement

---

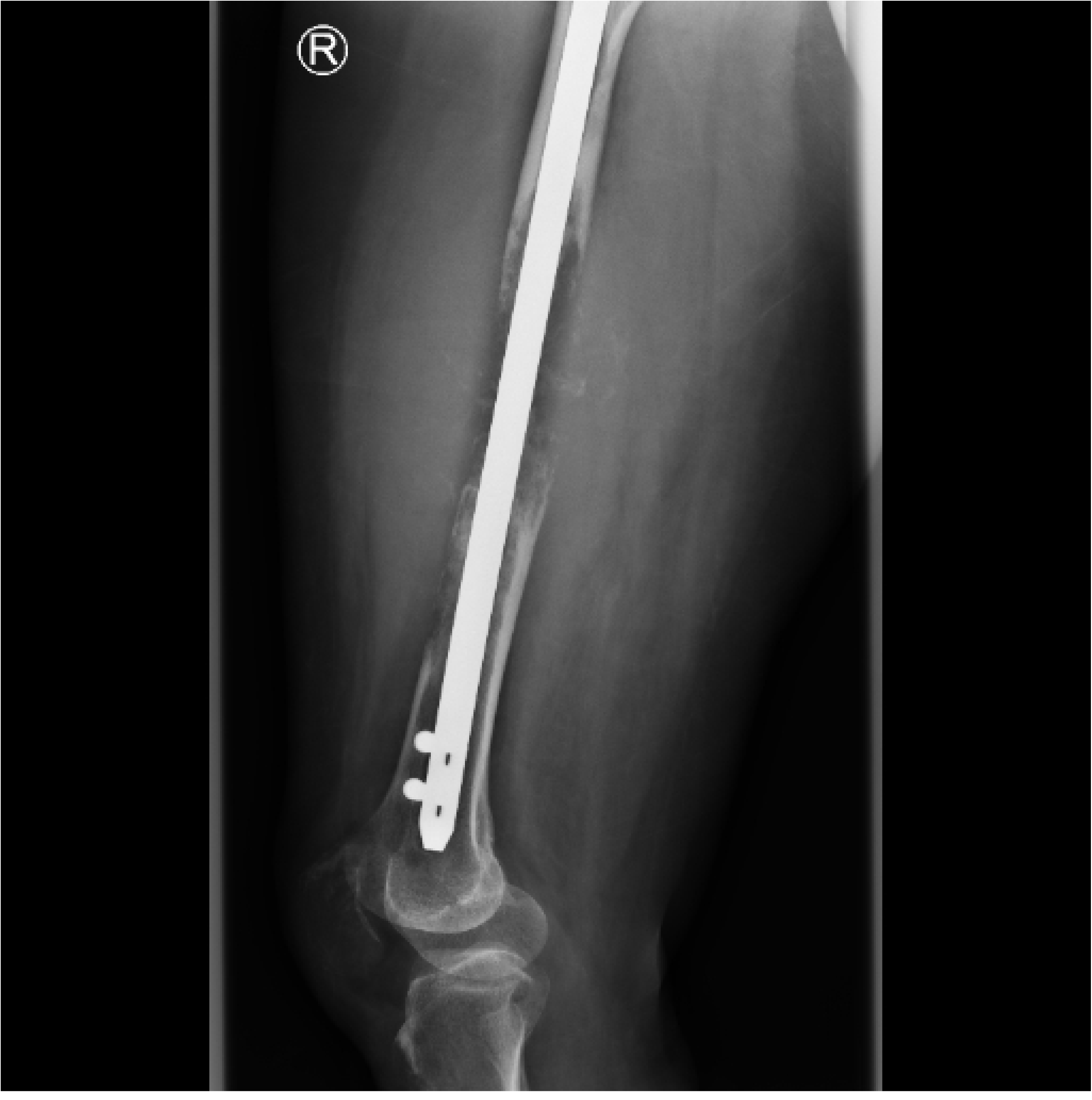

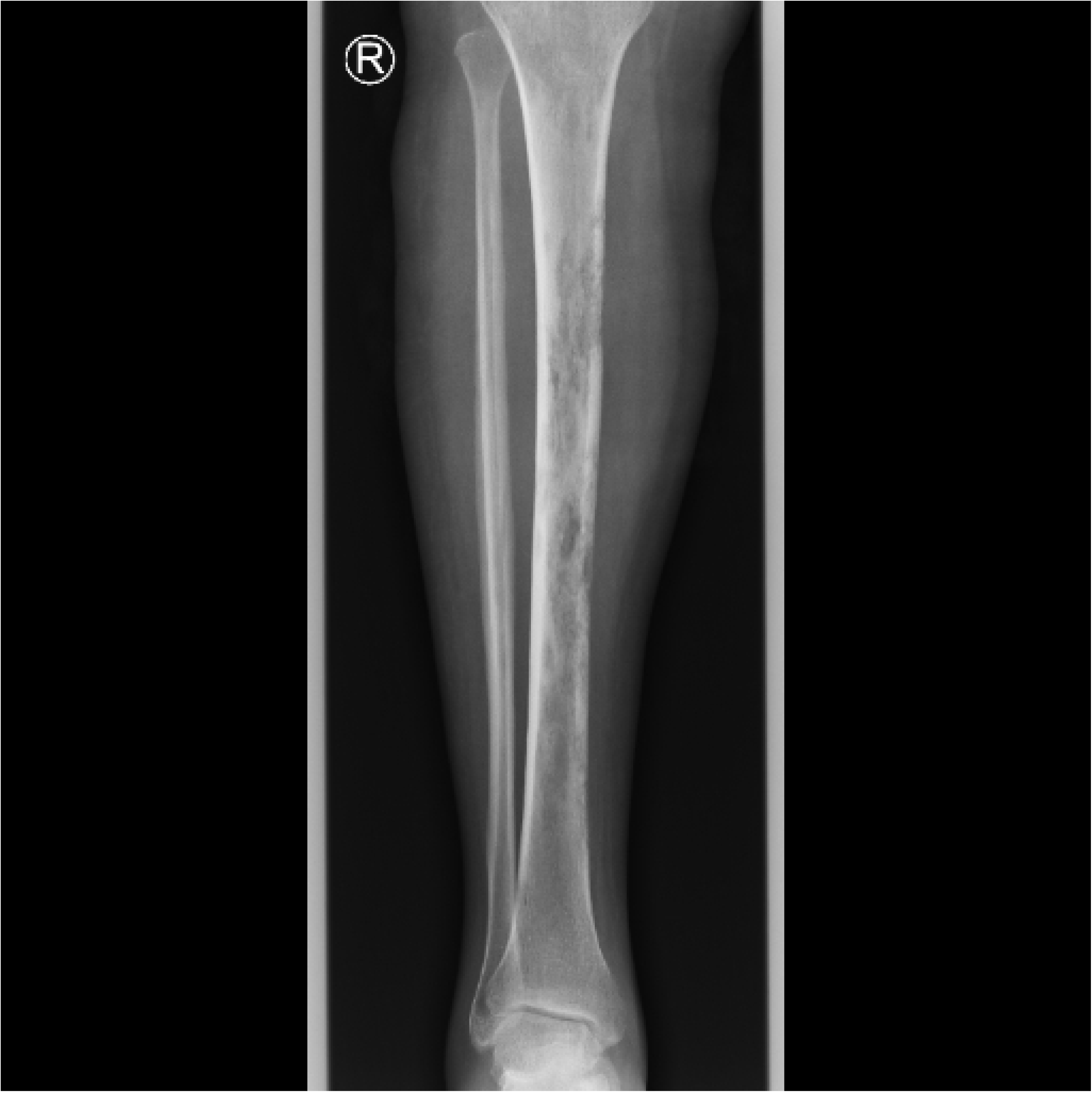

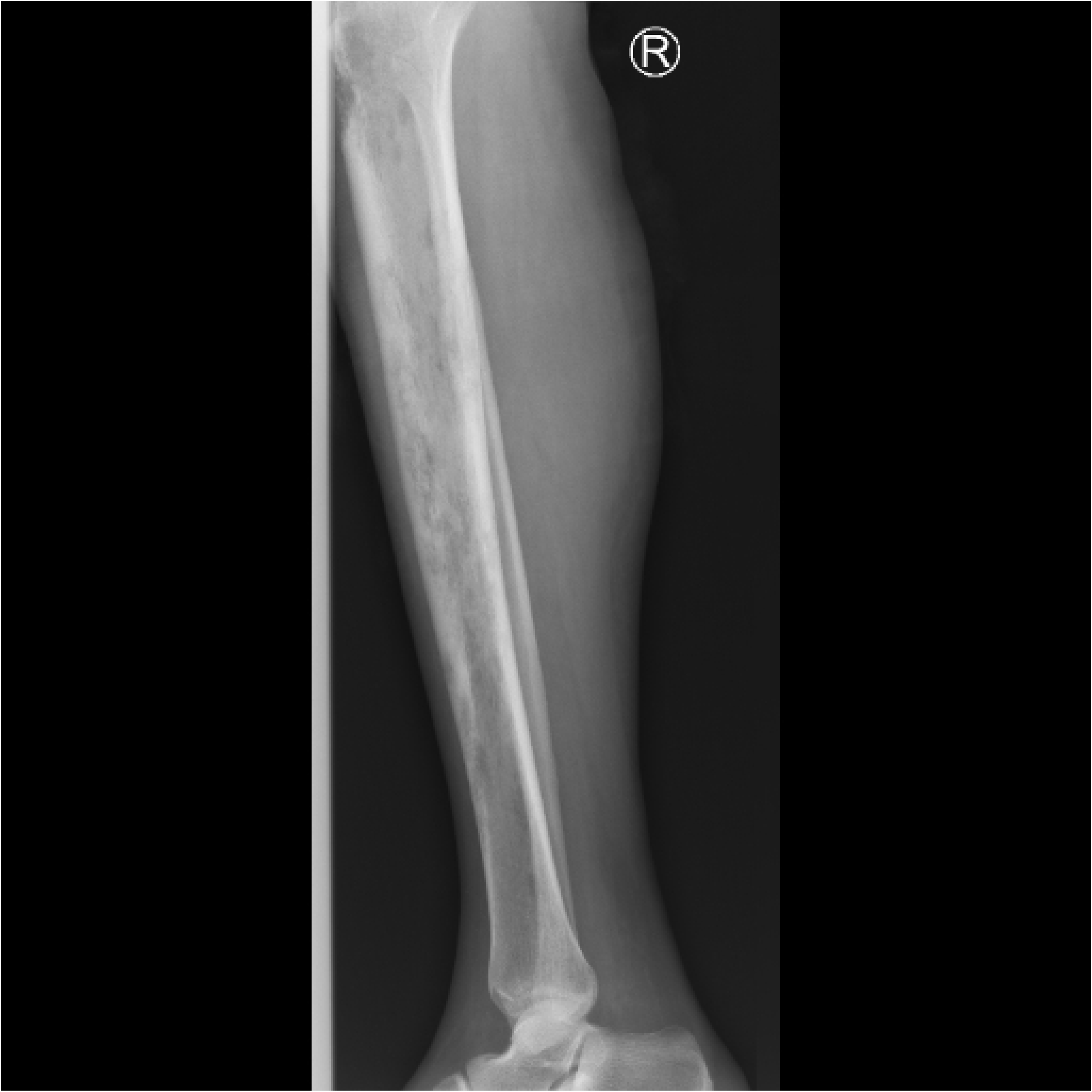

